# Projecting COVID-19 Cases and Subsequent Hospital Burden in Ohio

**DOI:** 10.1101/2022.07.27.22278117

**Authors:** Wasiur R. Khuda Bukhsh, Caleb Deen Bastian, Matthew Wascher, Colin Klaus, Saumya Yashmohini Sahai, Mark Weir, Eben Kenah, Elisabeth Root, Joseph H. Tien, Grzegorz Rempala

## Abstract

As the Coronavirus 2019 (COVID-19) disease started to spread rapidly in the state of Ohio, the Ecology, Epidemiology and Population Health (EEPH) program within the Infectious Diseases Institute (IDI) at the Ohio State University (OSU) took the initiative to offer epidemic modeling and decision analytics support to the Ohio Department of Health (ODH). This paper describes the methodology used by the OSU/IDI response modeling team to predict statewide cases of new infections as well as potential hospital burden in the state.

The methodology has two components: 1) A Dynamic Survival Analysis (DSA)-based statistical method to perform parameter inference, statewide prediction and uncertainty quantification. 2) A geographic component that down-projects statewide predicted counts to potential hospital burden across the state. We demonstrate the overall methodology with publicly available data. A Python implementation of the methodology has been made available publicly.

**Highlights:**

- We present a novel statistical approach called Dynamic Survival Analysis (DSA) to model an epidemic curve with incomplete data. The DSA approach is advantageous over standard statistical methods primarily because it does not require prior knowledge of the size of the susceptible population, the overall prevalence of the disease, and also the shape of the epidemic curve.
- The principal motivation behind the study was to obtain predictions of case counts of COVID-19 and the resulting hospital burden in the state of Ohio during the early phase of the pandemic.
- The proposed methodology was applied to the COVID-19 incidence data in the state of Ohio to support the Ohio Department of Health (ODH) and the Ohio Hospital Association (OHA) with predictions of hospital burden in each of the Hospital Catchment Areas (HCAs) of the state.

## 1. Introduction

The coronavirus 2019 (COVID-19) disease has resulted in 267 million confirmed cases and 5 million deaths reported globally as of 8 December 2021 [10]. As evidenced by epidemics in many countries, particularly in Italy and Spain, COVID-19 patient case loads have the potential to overwhelm healthcare systems [14].

The Infectious Diseases Institute (IDI) at the Ohio State University working in conjunction with the College of Public Health (CPH), the Department of Mathematics, and the Sustainability Institute (SI) established a working relationship with the Ohio Department of Health (ODH) to act as a service to the State, beginning in 2018. Based on this initial collaborative relationship, the Ecology Epidemiology and Population Health (EEPH) program within IDI took the initiative to offer epidemic modeling and decision analytics support to ODH preparatory planning and response to the COVID-19 pandemic, specifically the Ohio epidemic.

Just as in any computational modeling effort, there are many approaches and methods to choose from to model emerging epidemics. Different methods are suited to different situations in terms of data, scenarios, conditions, and urgency. This is no different for modeling the COVID-19 pandemic, either nationally or for the state of Ohio. Modeling approaches include: compartmental models [11, 28, 35], statistical models [16], and agent-based simulations [13]. For the state of Ohio, the OSU-IDI group has approached the modeling challenge on two fronts by developing: 1) projected statewide estimates of a time series of COVID-19 incidence, and 2) a geographic component that transforms the output of the statewide model to hospital burden by county or “hospital catchment” area.

## 2. Methods

### 2.1. Generalization of Methods Used

The predictive statewide model for Ohio comprises a dynamic network model, where network edges (contacts between nodes in the network) can interact with each other [24], but it has three key distinguishing features:

1. The model considers a dynamic network where the edges can be deactivated over time—supporting social distancing impacts more accurately.
2. The law of large numbers yields a set of differential equations describing the disease process on a large network [17] without requiring simulation methods.
3. The solutions of these differential equations can be used to estimate model parameters using a principled statistical approach based on survival analysis. We write an explicit likelihood for the parameters given data on times of illness onset [19]. Critically, this allows for a more accurate quantification of the uncertainty in the model predictions.

Because of these features, the DSA approach retains the tractability of an analytic model while incorporating complex human networks to better represent social interactions and distancing.

The projected statewide model takes data on illness onset dates of confirmed cases as input and produces estimates of COVID-19 incidence (*i*.*e*., new cases) in Ohio over time. This output is not age-stratified, but the age distribution of the new cases is assumed to match the age distribution of confirmed cases when projecting the number of hospitalizations, which occurs in the next step of the model.

This is done separately for illness severity and hospitalization since these rates vary for COVID-19 patients based on age and comorbidity stratification [31, 33]. Rather, the projected statewide model provides an estimate of the number of new and cumulative cases over time across the entirety of Ohio.

Estimates of the number of hospitalized COVID-19 cases are derived in the geographic component of the model. Because illness severity and the risk of hospitalization for COVID-19 patients vary according to age and comorbidities [31, 33], we use age structure and population density to distribute case counts from the statewide epidemic model across smaller geographic areas within the state. Within each geographic unit, we use its own age distribution to project hospitalization rates over time. Consequently, counties or hospital catchment areas that have a different demographic structure (*e*.*g*., older or younger populations) will differ in their COVID-19 hospitalizations over time.

### 2.2. Detailed Methods of the Predictive Model

There are challenges to using traditional compartmental models [25] to estimate future COVID-19 incidence. One of the most fundamental challenges is that these methods require knowledge of the size of the susceptible population. In our predictive model, we use an approach called *Dynamic Survival Analysis* (DSA) [20, 30, 34], which is an extension of *survival dynamical systems* [6, 19]. Details on this approach and the model development can be found in Appendix A.

Three key strengths of the DSA approach for predicting novel virus epidemics such as COVID-19 are:

- It does not require knowledge of the size of the susceptible population,
- It does not require information on overall disease prevalence in the population.
- It does not require prior knowledge of the shape of the epidemic curve.

Since testing has focused on the most symptomatic and severe cases, we have almost no information on the number of asymptomatic infections or those with less severe symptoms who are not tested. Because of the novel nature of this virus and the resulting pandemic, analysis of the epidemic curve cannot be based on previous epidemics caused by other viruses *e*.*g*. SARS-CoV-1, or influenzas. Because it relies on illness onset times rather than counts of new cases, the DSA method can incorporate a partial epidemic curve like the one produced by testing for COVID-19 in Ohio, which is affected by both limited testing capacity and undetected asymptomatic infections. The DSA method has been developed to handle such limited data in a manner that allows accurate quantification of uncertainty. The method is strengthened by its simplicity as it requires only a single differential equation. Parameters for the model are inferred using empirical temporal data on new illnesses, and the target output is a time series of expected future illnesses.

Initially, we used maximum likelihood estimates (MLEs) to fit the DSA model to Ohio data. However, as a substantial amount of data became available, we adopted a Hamiltonian Monte Carlo-based Bayesian approach to parameter inference and uncertainty quantification. An implementation of our inference method in the Python programming language is made available as a GitHub repository [4].

Our projected statewide model provides robust estimates of cases over time given partially observed daily counts of new illnesses. This is ideal in a setting where testing capacity is limited or changing due to constraints on lab capacity or detection limits. The counts of new illnesses are often known as an observed *epidemic curve* in the literature [36]. Our approach is derived from the general stochastic model of a pathogen spread across a contact network where the nodes represent individuals in a community [17]. As a working model of a contact network we use a type of random graph called a dynamic configuration model (CM) [7]. The mean-field approximation for epidemic processes on such networks is sometimes referred to as a pairwise model [21].

#### Assumptions

All models have specific assumptions used to develop and implement them. First, we assume that each individual in the network (node) has a number of neighbors (their degree). Subsequently, a local Markovian infectious pressure changes their status from *susceptible* (*S*) to *infective* (*I*) to *removed* (*R*). We further assume that the *R* individuals are no longer able to transmit the infection and cannot be reinfected—an unknown, but readily assumed component of contemporary models. For the dynamics of the network model, individuals transition between the *S, I*, and *R* compartments based on the following principles that allow the extraction of a mathematical model of the ongoing epidemic based on observable data:

1. Disease spread occurs over a network of contacts, that is, an infectious individual can infect their immediate neighbors at a fixed positive rate. It is implicitly assumed that the average number of a person’s contacts is also positive.
2. Each infected individual recovers from infection at a positive rate, **or** is restricted from contacting their network neighbors through mandatory or voluntary isolation at a positive rate.
3. Once infected, an infected individual has an infectious period that is assumed to follow an exponential distribution.
4. People who are ill remain infectious, and a partial count of new illnesses is observed over time with a negligible chance of misdiagnosis (*i*.*e*., false positives).

#### Estimating Transmission Rates

The complete model description and detailed development can be found in the appendix. In brief, the statistical model is used to estimate the number of people and timing of transfer between states *S, I*, and *R*. Then through substitution, a term *S*_*t*_, the number of susceptible people at time *t* is developed. Embedded within this *S*_*t*_ is an improper survival function for the *time to the onset of illness in a randomly chosen susceptible person*. Using this survival function approach, the time to infection of a randomly selected susceptible person within a large population follows a temporal pattern determined by probability laws. After we estimate the time series of susceptibles, we can estimate the probability of a randomly selected susceptible individual being infected *during the lifetime of an epidemic*. From this we develop the conditional probability of remaining susceptible past time *t*.

#### Estimating Dropout and Recovery Rates

The impact of social distancing is accomplished via a generalized approach to a person being dropped from the network. As has been visualized elsewhere, when a person is removed from the network, their neighbors and contacts within the network are removed, which limits transmission. This is accomplished by estimating the rate of infectious contact within a network (*β* in the appendix), and then considering drop outs as a function of recovery rate (details in appendix).

#### Changes in Parameters Due to Interventions

Introducing and then easing restrictions on social distancing in the state has likely resulted in changes in parameters. The effect of such changes is incorporated into the predictive model.

#### 2.2.1. Estimating Hospitalization and ICU Admission Onset

After the statewide model produces time series estimates of cases across the state, these are then translated into estimates of hospitalizations and subsequent ICU admission. The complete derivation of this model can be seen in Appendix B. In short, this is developed in three steps and is integrated with the geographic modeling methods described next.

1. Estimate case onset for each age group based on age stratification in each geographic area modeled.
2. Estimate the number of severe cases that will need hospitalization based first on CDC data [32] and then updated with Ohio-specific data as those data become more conducive to predictive modeling.
3. Estimate a probability distribution (details in Appendix B) for the time from case onset to hospitalization as a function of age, sex, and race. Using the same variables, we also estimated the time from hospitalization to discharge or death and the probability of being admitted to the ICU. This provides a means to model hospital occupancy based on Ohio-specific data.

### 2.3. Geographic Modeling Methods

The time series of estimated cases of COVID-19 illness are output from the statewide model described above. These outputs are then used to assess hospital burden (*eg*. hospitalizations and ICU admissions) using age structure of the population and the estimated length of time a patient will be in a hospital or ICU bed. This involved a three step estimation process:

1. The proportion of the total Ohio population residing in each geographic area (*e.g*., county or hospital catchment area) was estimated from U.S. Census data. Daily case counts were distributed across geographic areas using this proportion, essentially distributing cases by population density. This created a time series of projected case counts by geographic area.
2. To account for age differentials in hospitalization and ICU use noted by both the popular press and in the scientific literature, we estimated the proportion of new cases expected to require hospitalization or admission to the ICU using the age distribution in each geographic unit at each time step. Initially, we used estimates from the Morbidity and Mortality Weekly Report (MMWR) published by CDC [32]. These nationwide estimates of the age distribution of hospitalization were used only while the number of hospitalizations and ICU admissions in Ohio remained too low to make reliable forecasts. Ohio rates were substituted in subsequent runs of the model. We estimated the number of hospitalized cases by assuming higher rates of hospitalization for older segments of the population. Thus, in counties with an older age structure, a larger proportion of cases would convert into a hospitalization.
3. We used the estimated number of new hospitalizations and the average length of stay (LOS) to estimate the total hospital burden for each day in the time series. Initially, the average LOS was derived from the literature, but we used observed LOS from Ohio hospitals when enough COVID-19 cases were identified to provide robust estimates. We used a bi-modal distribution, with an average LOS of 5 days for non-ICU patients and 14 days for those requiring ICU care. For each time step, the number of new hospitalized cases estimated from the model was added to the number of cases still in the hospital. The cases that “timed out” of their hospital stay due to the LOS parameter were subtracted from the total. This created an estimated net number of patients in the hospital for each day in the time series which accounted for “patient stacking” over time.

Once daily hospital counts were estimated, we compared these to the reported number of COVID available beds pooled across all hospitals in a geographic area. This allowed us to understand when and where hospital bed need might exceed current capacity.

#### 2.3.1. Description of Data

##### Demographic data

Demographic data on the age distribution for Ohio counties and ZIP Codes were obtained from the U.S. Census Bureau’s 5-year American Community Survey 2014-2018 estimates [8]. The ACS data were used because the sample size was large enough for small geographies for reasonable standard errors and stable estimates. The Census Bureau does not develop data products for the United States Postal Service (USPS) ZIP Codes. Rather, ZIP Code Tabulation Areas (ZCTAs) are generalized areal representations of USPS ZIP Code service areas. We mapped the Census ZCTAs to ZIP Codes and used population estimates for ZCTAs. We used table B01001 that breaks out population counts by sex and 5 year age groups. Figure 1 shows the distribuion of the high risk population (age 55+) by county and Hospital Catchment Area in Ohio.

**Figure 1:**
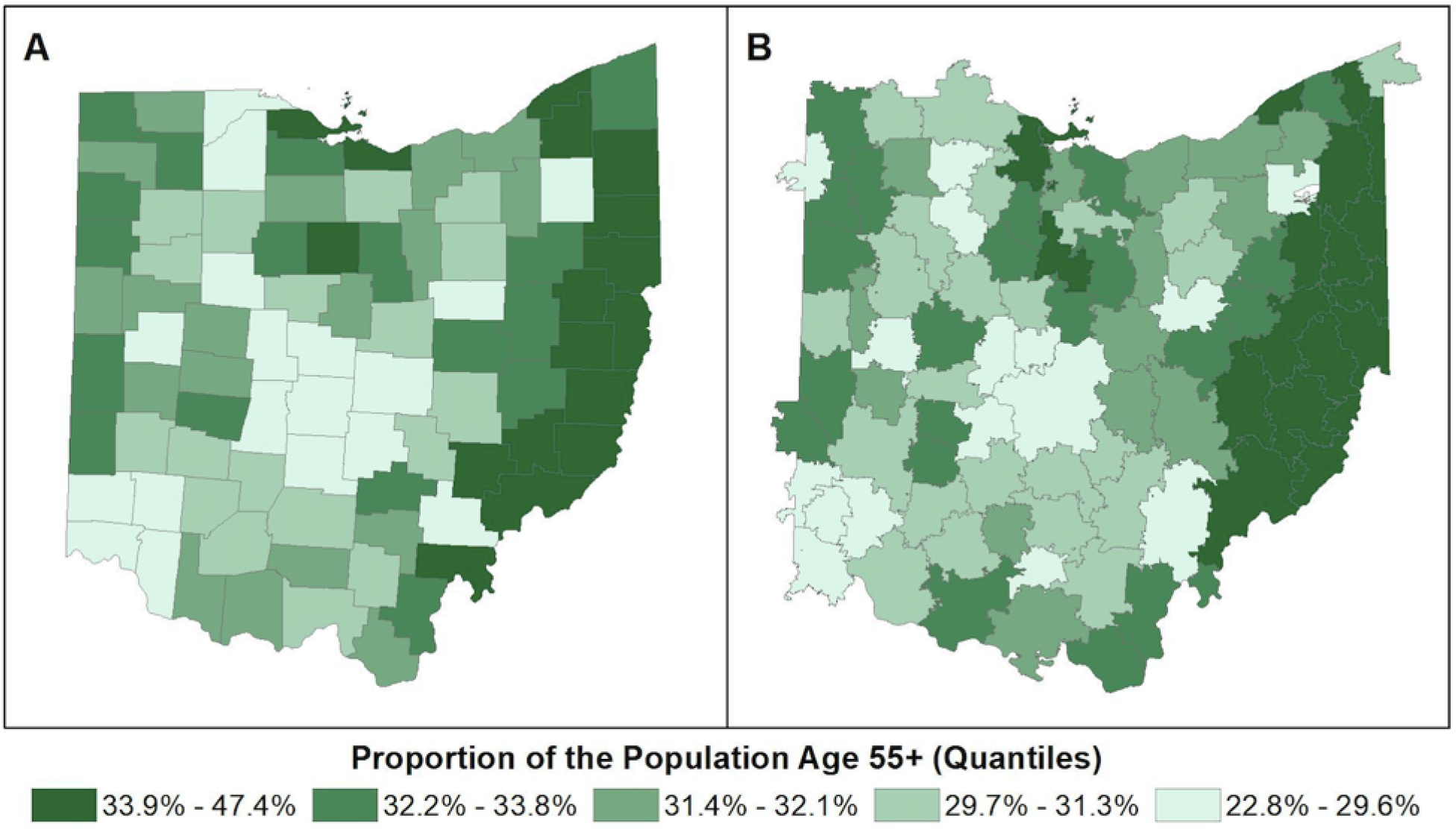
Proportion of the population over the age of 55 in A) counties and B) hospital catchment areas

##### OHA Hospital and Bed Data

The Ohio Hospital Association (OHA) provided data on the location of all hospitals in the state and the number of registered beds for each facility. Beds were broken out into several categories: Airborne Isolation, Critical Care, General Medical/Surgical Beds, and Extracorporeal Membrane Oxygenation (ECMO) beds. These bed types represent two levels of care required for COVID-19 patients, general hospital care for severe disease, and ICU beds for patients requiring ventilation. The OHA data contains information that constitutes a “trade secret” under Ohio Revised Code Section 1333.61.

##### Description of OHA hospital market share data

OHA provided data on hospital market share derived from administrative hospital claims from the past year (January 2019-December 2019, inclusive). For each hospital, patient encounters were grouped by patient zip code. A hospital’s market area included zip codes that represented the top 80% of all encounters at that hospital.

#### 2.3.2. Definition of Hospital Catchment Areas

We developed small area estimates from state-level model predictions for counties and hospital catchment areas (HCA). Boundary files for the 88 Ohio counties were obtained from the U.S. Census Cartographic Boundary Files [9]. We developed hospital catchment areas using an approach modified from the Dartmouth Atlas Project [1]. We define a hospital catchment area as a collection of ZIP codes whose residents receive most of their hospital care from the hospital(s) in that region. Figure 1-B shows the HCAs developed and mapped. Note that in figure 1-B there are HCAs that cross over into neighboring states. This is explained in more detail in the subsection below.

HCA definitions depend on the integrated use of geospatial methods that grouped each zip code in the state with the most geographically proximate hospital and modified these groupings using data on hospital market share by zip code. Only hospitals with acute care beds that could be used for COVID-19 patients were included in the analysis. We excluded facilities such as long-term acute care (LTAC), hospice, orthopedic, rehabilitation or psychiatric/behavioral health hospitals and freestanding ERs. We defined HCAs using three steps:

1. The location of all hospitals in Ohio, and those in Michigan, Indiana, West Virginia, Kentucky and Pennsylvania located on the border with Ohio, was used to generate a Voronoi diagram with hospitals as generating points [26]. We included hospital in neighboring states in the Voronoi analysis to avoid edge effects, which would attribute patients to Ohio hospitals that typically use hospitals in other states. This yielded 233 distinct areas, one for each hospital generator. Twenty-eight areas were subsequently deleted because they included no area within OH.
2. Voronoi polygons were overlaid with ZIP codes. ZIP codes were assigned to the Voronoi polygon if their centroid fell within the polygon. This ensured that each ZIP code was associated with the most geographically proximate hospital and divided the state into groupings of zip codes assigned to each hospital.
3. Using the OHA hospital market share file, we examined the level of agreement between each Voronoi polygon and market share zip codes for each hospital. In cases where adjacent Voronoi polygons were generated by hospitals that also shared 60% or more of their market share zip codes, we aggregated these polygons to create one hospital catchment area. In large metropolitan areas with many hospitals, we aggregated groupings of Voronoi polygons to create regional catchment areas. In rural areas, this typically resulted in aggregating two adjacent polygons.

Using this procedure, we generated 96 Hospital Catchment Areas for the state of Ohio. Some HCAs included zip codes from neighboring states, and some Ohio ZIP codes were included in HCAs for non-Ohio hospitals. HCAs are shown in Figure 2.

**Figure 2:**
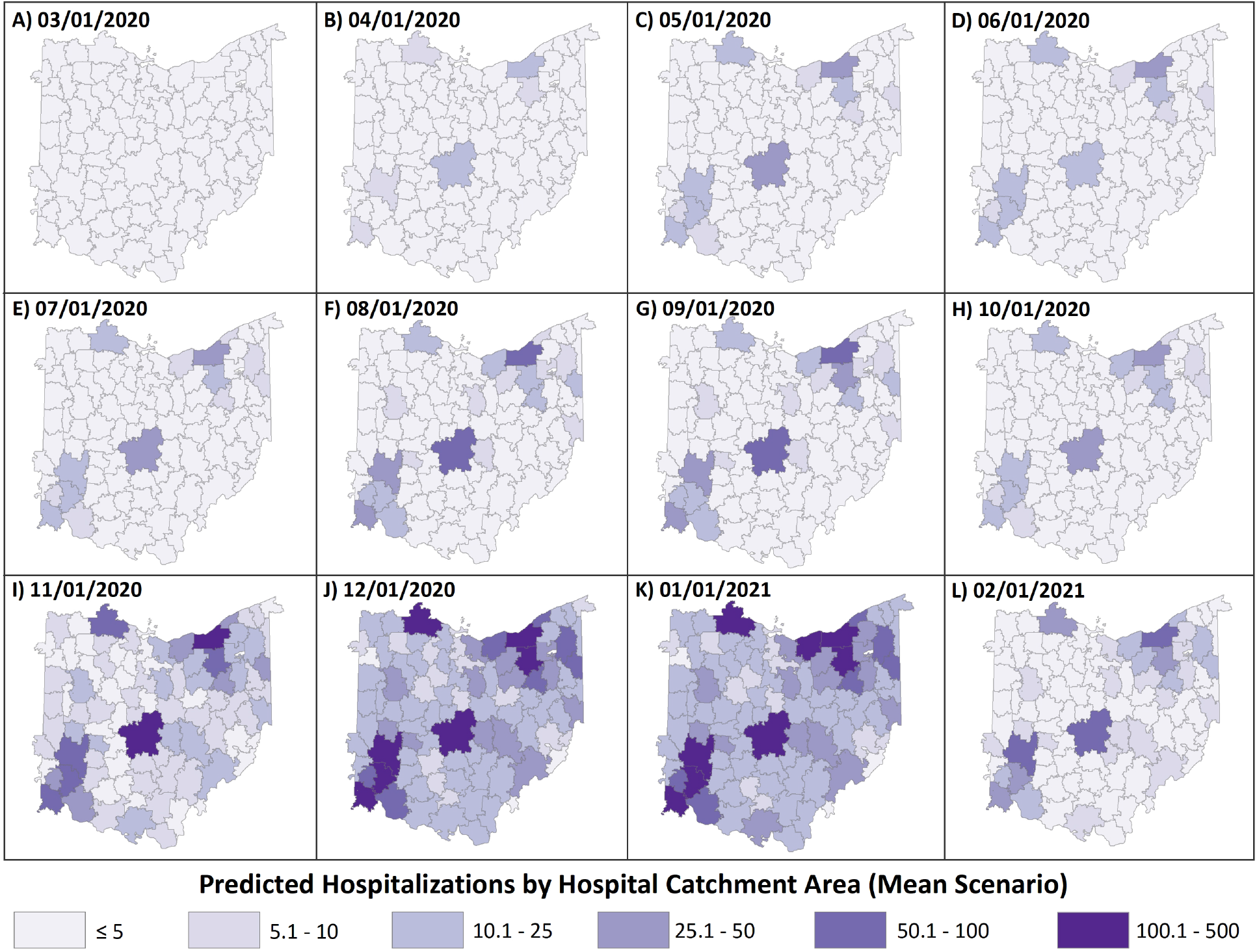
Estimates of hospital burden mapped to hospital catchment areas for 12 days during the pandemic

## 3. Results

Here, we present a brief summary of our model fits and predictions. One of the hallmarks of the COVID-19 pandemic has been the change in human bahavior due to various interventions throughout the course of the pandemic. As a consequence, the COVID-19 epidemic curve in Ohio deviated much from a typical epidemic curve. The Ohio curve initially followed an exponential growth phase, then a phase of steady linear growth and a decline, and then finally, due to reopening of the state, another phase of exponential growth. As such, it is natural to believe the parameters are potentially different in these different phases. Therefore, we fit the DSA model with multiple change points.

In Figure 3, we compare actual counts of daily new infections against the model predictions. As one can see, the fitted trajectories follow the observed epidemic curve reasonably well. The inner confidence bound is the true posterior confidence bound obtained point-wise. However, it is worth noting that predicted trajectories underestimate the variance or over-dispersion in the observed counts of daily new infections. Therefore, we adopted an empirical variance adjustment method to account for this possible underestimation. The broader confidence bound corresponds to the variance adjusted trajectories. More details on the variance adjustment method and other diagnostic plots are provided in Appendix B. The daily counts thus predicted are then down-projected into predictions of hospitalization surge across the state in the geographic component of the model.

**Figure 3:**
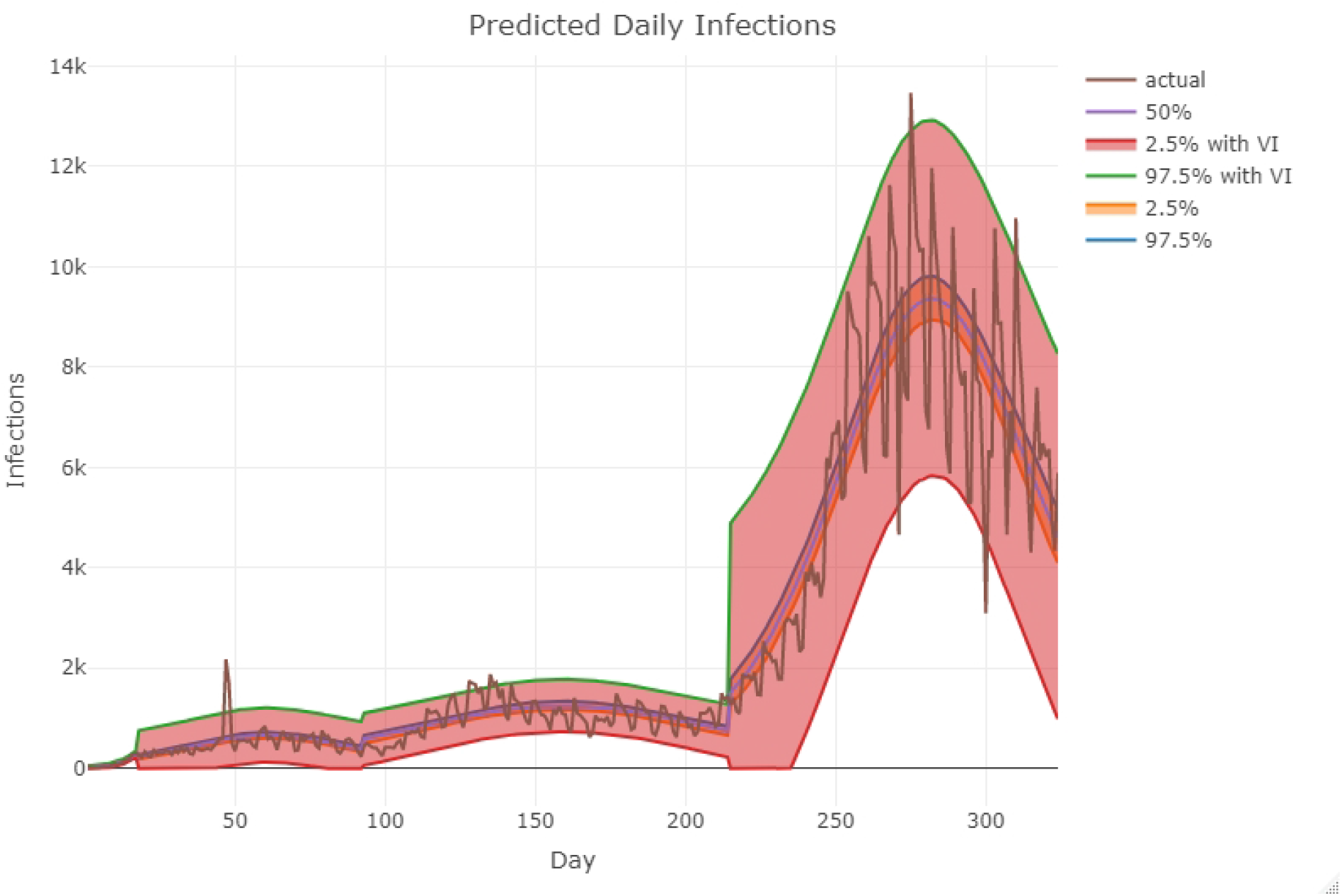
Predictions based on the fitted model vs actual daily incidence in Ohio as reported by the Ohio Department of Health. We have assumed three change points here corresponding to four density segments. Day zero corresponds to March 1, 2020. The first change point is on March 17, 2020. The second change point is on June 1, 2020 after which the state observed another phase of exponential growth. The third change point is on October 1, 2021, which marks the beginning of second wave characterized by a severe exponential growth phase. The broader (outer) confidence bound corresponds to the variance adjusted trajectories.

## 4. Discussion

### 4.1. Comparison with Other Approaches

Although, due to the significant differences in approach, it is difficult to directly compare our projective model to other contemporary methods, some indirect comparisons may be offered. The susceptible-infectious-recovered (SIR) framework is the basis for many COVID-19 epidemic models [11, 22, 28, 35]. Several websites provide implementations of the SIR framework and present scenarios with different interventions such as social distancing affecting the transmission parameter *β* [11, 28], thus allowing for different hypothetical scenarios to be explored. A key difference, however, is that phase-space variables from the SIR model are infections, rather than symptomatic cases. Consequently, knowledge of asymptomatic to symptomatic infections is needed to translate output from these models to cases and hospitalizations. Recent work [11] has attempted to extend the basic SIR model to include additional compartments corresponding to incubation, presymptomatic infectious, stages of symptomatic infectious, and hospitalization. Considering the lack of clear information on these types of outcomes, it is clearly difficult to implement this model as such detailed data are not currently available.

The current SIR tools [11, 28] are suited for scenario exploration, like illustrating the *flattening of the curve* under different levels of social distancing compliance. Unfortunately, they are less suited for forecasts as model parameters are not calibrated based upon case or outcome data. A recent attempt at taking a meta-population approach with SEIR dynamics in each patch, to estimate model parameters (including proportion of asymptomatic infection) based upon case counts from China was given in [22]. This resulted in a spatially explicit model without age-structure. Conversely, [35] is an example of a model with age structure but not spatial: Weitz extends the SEIR framework to include age groups, and estimates model parameters based upon hospitalization and death counts from Georgia. Further work from the University of Washington uses mortality data as model inputs, but takes a purely statistical approach in fitting a sigmoidal curve to cumulative COVID-19 deaths using a mixed-effects model [16]. Another alternative is an agent-based model such as that used in [13]. Similar to [22], [13] forecast a very large number of infections with COVID-19. Considering the level of uncertainty in the data being input into all of these models, we rationalize that a model output of a time series of illnesses is the more reasonable approach. Similarly, using the simplified framework and structure of our approach allow for greater flexibility under uncertain data inputs into the model, and a lack of typically required information such as total susceptible population, or asymptomatic rates.

## Data Availability

All data that produced in the present study that are not considered a "trade secret" are available upon reasonable request to the authors or from the site https://coronavirus.ohio.gov/dashboards/overview

## Acknowledgment

We would like to acknowledge Dominick Winecki for developing useful shell scripts during the early hectic days of the pandemic. We also acknowledge Ian Dunn for his help with the GIS software and graphics.

## Funding

WKB acknowledges the President’s Postdoctoral Scholars Program (PPSP) of the Ohio State University (OSU). The work of GAR and EK was partially funded by the National Science Foundation (NSF) Rapid Grant DMS-2027001. EK and WKB were partially funded by National Institute of Allergy and Infectious Disease (NIAID) grant R01 AI116770. The content is solely the responsibility of the authors and does not represent the official views of the NSF or NIAID.

## Data Availability & Software

The software to perform model fit along with a data example is available from [4]. The onset data used in this paper can be downloaded from the Ohio Department of Health (ODH) COVID-19 dashboard at https://coronavirus.ohio.gov/dashboards/overview. Probability distributions for the time from disease onset to hospitalization, length of stay, and probability of admission to the ICU were based on analysis of non-public data from ODH.

## Additional Materials

A video presentation describing the methodology used for epidemic size predictions is available from [29].

### A. Predicting Statewide Cases of COVID-19

#### A.1. Detailed Description of Project Mode Development

This derivation is taken in whole from the manuscript in preparation [5]. We will outline the derivation of a simple but powerful general modeling framework that provides robust estimates of the quantities relevant to monitoring local outbreaks where only limited amount of information is available through partially observed daily counts of new (symptomatic) infections - *e.g. illnesses*. Our approach is derived from the general stochastic model of a contagion spread across a contact network of nodes representing individuals in a community [17]. As a working model of a contact network we use a dynamic configuration model (CM)-type random graph (see, *e.g*.,[7]). Such a model is often also referred to as a *pairwise model* [21].

Briefly, we assume that each node has its degree and that the nodes may change their status from the initial “Susceptible” (*S*) to “Infective” (*I*) (or “Infectious”) and, finally, to “Removed” (*R*), according to their local Markovian infectious pressure (hazard). We assume that the “Removed” individuals are no longer able to pass infection and cannot be reinfected.

#### A.2. Network Dynamics Assumptions

The dynamic model of individuals transition between the states *S, I* and *R* is based on several simple principles that allow to extract a mathematical model of the ongoing epidemic and relate it to observable data:

1. The spread occurs over a network of contacts, that is, an infectious individual may only infect his/her immediate neighbors at fixed rate *β* > 0; it is assumed that the average number of node’s contacts is *μ* > 0.
2. The infected individual may recover at rate *γ* > 0 or be restricted from contacting his/her network neighbors either through mandatory or voluntary quarantine at rate *δ* > 0.
3. The infected individuals stay infected for a random amount of time distributed according to an exponential distribution.
4. The symptomatic infectives are infectious and the partial count of new infectives is observed over time with a negligible chance of misdiagnosis (false positives).

We note that the model as described here extends previous work studying epidemics on CM-random networks (e.g., [23]) and that this formulation above can also account for extensions of the basic SIR compartments to include a latent period (up to 14 days for COVID-19, see [25]), as well as more general staged progression models [15].

Instead of directly analyzing the stochastic CM model described above, which is challenging due to the heterogeneity in the number of contacts and the evolution of the connectivity structure (e.g., [3, 18]), we will make use of the general results on the mean field approximation [2, 12], and the convergence of the random infection hazard in large networks. Specifically, as shown in [17] under the assumption of the Poisson-type degree distribution (for instance the node degree is Poisson or negative-binomial distributed), the mean field approximation of the dynamics is given by the following set of differential equations (dots denote time derivatives)

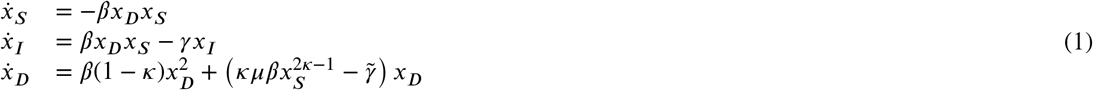

where the pair (*x*_*S*_, *x*_*I*_) describes the relative number of susceptibles and infecteds, *x*_*D*_ = *x*_*SI*_ /*x*_*S*_ is the relative density of infectious connections, *κ* is the average contact network density^1^, and 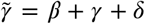. The usual initial conditions are *x*_*S*_ (0) = 1; *x*_*I*_ (0) = *ρ* > 0; *x*_*D*_(0) = *μρ*.

#### A.3. Estimating Reproduction Rate

For the purpose of statistical analysis of the system (1) we make an assumption that only the empirical counts of the new infected are available in practice. Dividing last equation in (1) by the first one, solving for *x*_*D*_ in terms of *x*_*S*_ and substituting back into the first equation we obtain the reduced system with only one equation describing the decay of susceptibles. To simplify notation, denote *S*_*t*_ := *x*_*S*_ (*t*) to obtain

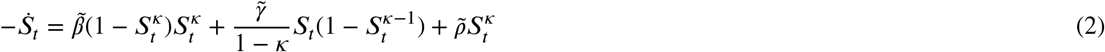

where *S*_0_ = 1 and 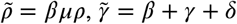, and 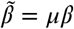.

Note that the equation (2) is defined for *κ* = 1 by taking the limit, *κ* → 1. The value of the basic reproduction number *R*_0_ for both reduced (2) and full (1) system is

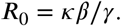

The condition *κ* = 1 implies the Poisson degree assumption for the pairwise model and reduces (2) to

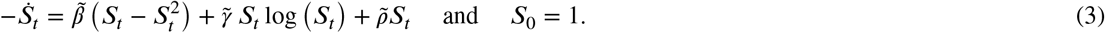

Instead of thinking about *S*_*t*_ as a proportion of susceptibles it is convenient to think about *S*_*t*_ as an improper survival function of *the time to infection* of a single randomly chosen susceptible. Then *S*_*t*_ has an improper density 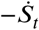 (see [19]). It is improper since 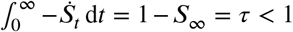 where *τ* is defined below (see also [6, Example 2]). Under this survival function interpretation, the infection time for a randomly selected initially susceptible individual (in an infinite population) follows a temporal pattern according to the probability law *S*_*t*_ given by (2) or (3). When we observe only a partial epidemic trajectory, say until time *T*, then the observed infection time is conditional on the infection occurring by time *T*, that is, on an event that has probability *τ*_*T*_ = 1 − *S*_*T*_. It is easy to show that as *T* → ∞ then *τ*_*T*_ → *τ*_∞_ = *τ*, the probability of a randomly selected susceptible individual *being infected during the lifetime of an epidemic*. (One may think of *τ* also as the final proportion of infected in the epidemic in infinite population). Then the conditional density of symptom onset is

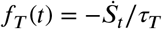

which is simply the scaled derivative of the probability of staying susceptible past time *t* (denoted *S*_*t*_). Accordingly, setting *θ* = (*κ, β, γ, ρ*), the approximate likelihood of the joint symptom times (epidemic curve) of *n* observed new cases by current time *T* in an infinite population (see [19]) is given by

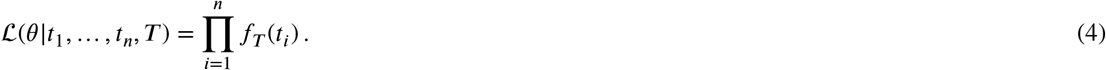

Although the expression above looks simple, note that the function *f*_*T*_ (*t*) depends upon the vector of parameters *θ* only implicitly through the differential equations (2) or (3).

#### A.4. Estimating Dropout and Recovery Rates

Recall 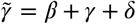. Given 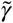, we may estimate the recovery rate *γ* from the recovery density. Then assuming *β* is negligible, we have the approximate expression for drop-out

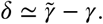

To estimate *γ* we consider now the recovery density. As shown in [19], the density of daily recovery times is given by

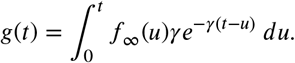

For practical model fitting, a shift parameter *ε* ∈ ℛ may be needed as

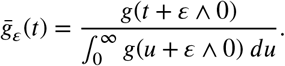

The overall density of recovery is then the following mixture

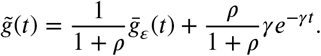

The analogous likelihood as in (4) can be now produced using the conditional recovery density

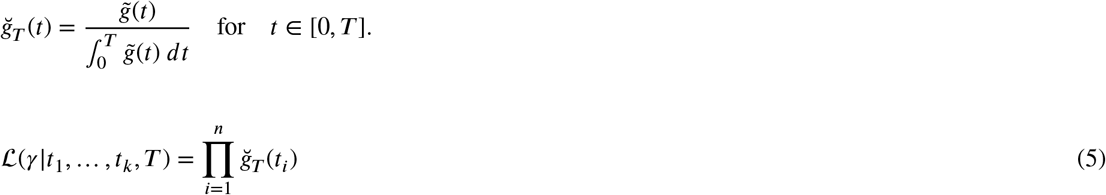

This likelihood is used to estimate *γ* directly conditional on estimated *S*_*t*_ (and thus *f*_∞_).

#### A.5. Incorporation of a change point

As outlined in an IDI blog post by the COVID-19 response modeling team [27], the initial exponential growth phase was followed by a plateau, a slow ascent, a decline, and then, a second exponential growth phase. This pattern can be attributed to changes in human behavior. As a consequence of the resultant unusual epidemic curve, we need to enhance the model to accommodate change points. In particular, we assume the parameter *θ* takes different values in different segments of the epidemic separated by the change points. For the sake of simplicity, we illustrate the revised likelihood function with a single change point.

Let *A* = (0, *T*^⋆^], and *B* = (*T*^⋆^, *T*] denote the two segments of the epidemic with a change point at *T*^⋆^ (*< T*). We assume the parameter *θ* takes value *θ*_*A*_ in segment *A* and *θ*_*B*_ in segment *B*. Then, the conditional densities in each of the segments are defined as

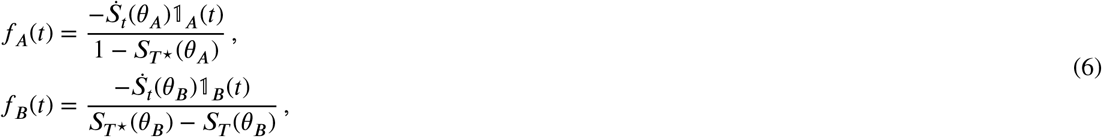

where *S*_*t*_(*θ*_*A*_) (or *S*_*t*_(*θ*_*B*_)) is the solution to either (2) or (3) with *θ* = *θ*_*A*_ (or *θ* = *θ*_*B*_). Write *τ*_*T*_ (·) = 1 − *S*_*T*_ (·). Then, the conditional density over the entire time interval [*θ, T*] is given by

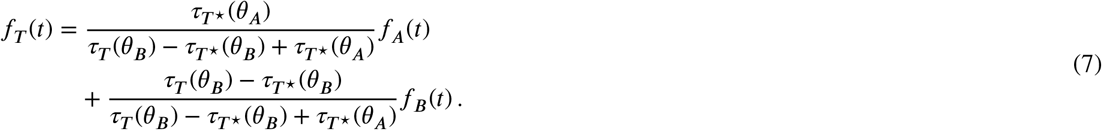

The final likelihood function looks exactly like (4) with the conditional density *f*_*T*_ given above in (7).

#### A.6. Estimating Size of an Outbreak

We assume that the size of an outbreak *k*_∞_ is a fixed integer representing the likely number of total infections in the contact network of the confirmed cases only. Hence *k*_∞_ is not the prevalence of the disease in the population but rather just an estimate of the total outbreak size in the community of *n* individuals where we see infections. We estimate *n* at any given time *T* by the discount estimator 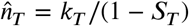 where *k*_*T*_ is the number of cases observed by time *T*. Then we estimate the total number of cases by the end of an epidemic as

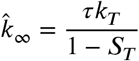

where *τ* = 1 − *S*_∞_ is the final probability of infection defined in Section 2.2.

#### A.7. Prediction and uncertainty quantification

We follow a Hamiltonian Monte Carlo-based Bayesian approach for prediction and uncertainty quantification. We assume (independent) non-informative priors for the parameters 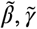 and 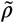. We approximate the posterior density of *θ* by drawing posterior samples using the *No U-Turn Sampling* (NUTS) method. As a point estimate of *θ*, we take the mean of the posterior density. The cumulative epidemic curve obtained as a solution to either (2) or (3) corresponding to the point estimate defined above gives us the most likely trajectory. The posterior samples of 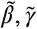 and 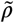 are used to generate a pointwise Monte Carlo confidence interval around the most likely trajectory. In other words, we essentially generate predicted trajectories corresponding to the posterior samples and then compute appropriate quantiles at desired time points to get the confidence interval. However, as shown in Figure 3, the confidence bound obtained this way is narrow and suggests that the procedure might be underestimating the variance or over-dispersion in the observed daily new case counts. This over-dispersion could be a result of testing delays or other systematic issues with data collection. Therefore, we adopt an empirical variance adjustment method.

In simple terms, the empirical variance adjustment could be explained as follows: We first smooth the observed counts using a kernel smoother with span = 0.2. We used the function `supersmoother` in R programming language. Then, we estimate the variance inflation factor by averaging the squared difference of the actual daily new case counts and the smoothed ones. Finally, the variance adjusted confidence bounds are obtained by multiplying the upper confidence bounds from the corresponding a Gaussian approximation with the standard deviation of the variance inflation factor and taking lower confidence bound from the DSA fits.

#### A.8. Additional numerical results

Here, we present additional diagnostic figures for our model fit. In Figure 4, we show the estimated density of the infection times in the three segments of the epidemic. This is the density that contributes to the likelihood function (4). The posterior densities of the fitted parameters in the three segments are shown in Figure 4. Finally, as a diagnostic measure of the convergence of the Hamiltonian Monte Carlo chains, we show the trace plots of the fitted parameters in Figure 7.

**Figure 4:**
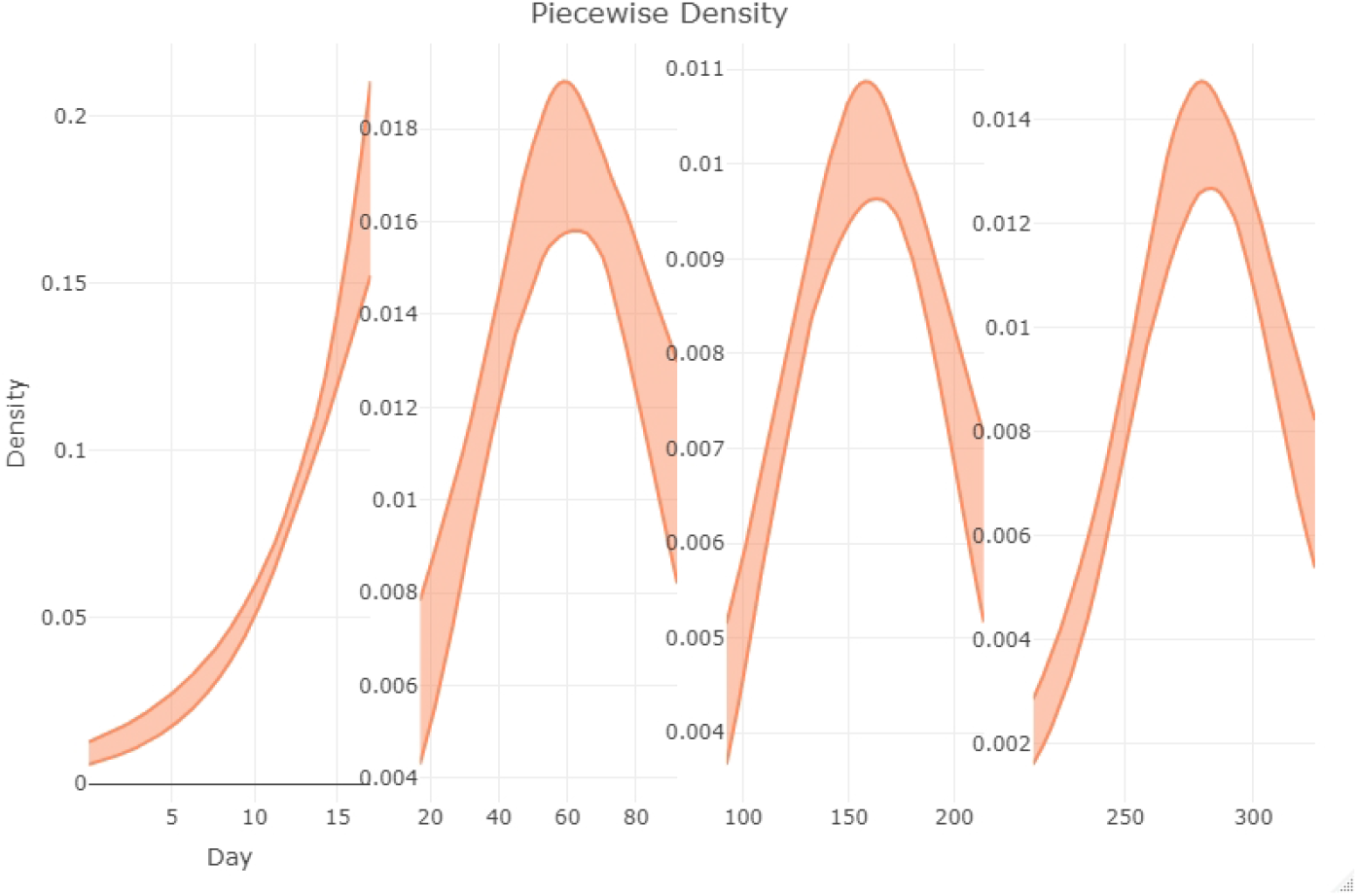
Estimated density of infection times in the four segments of the epidemic.

**Figure 5:**
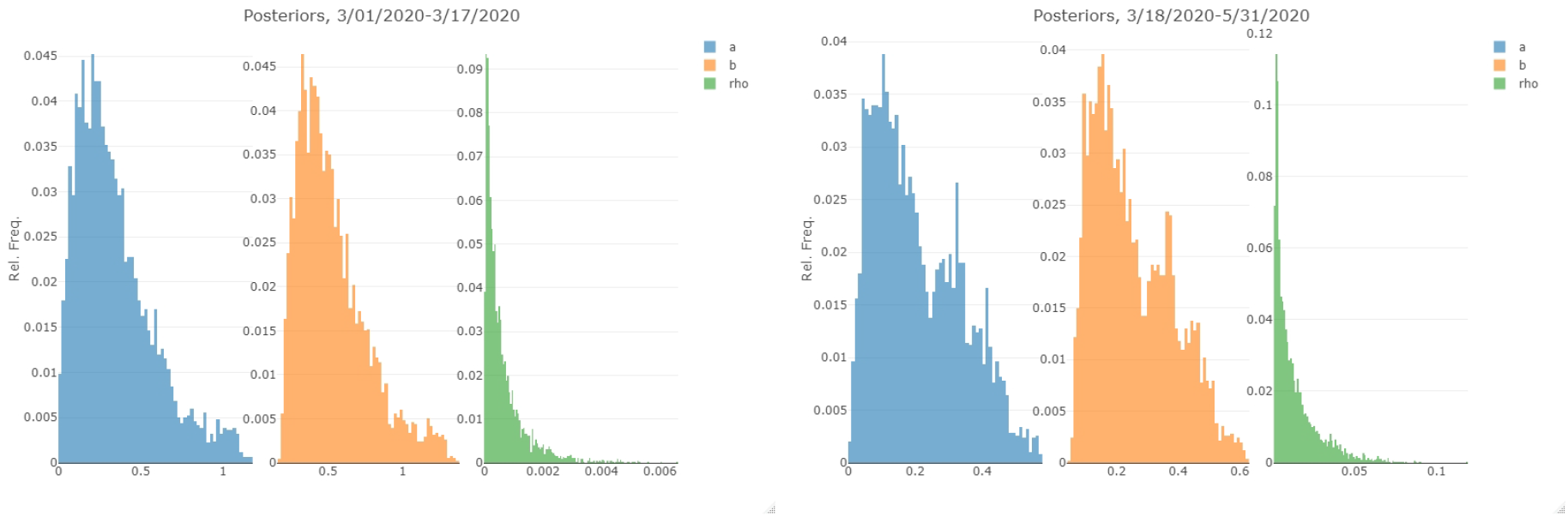
Posterior densities of the fitted parameters in the first two segments of the pandemic.

**Figure 6:**
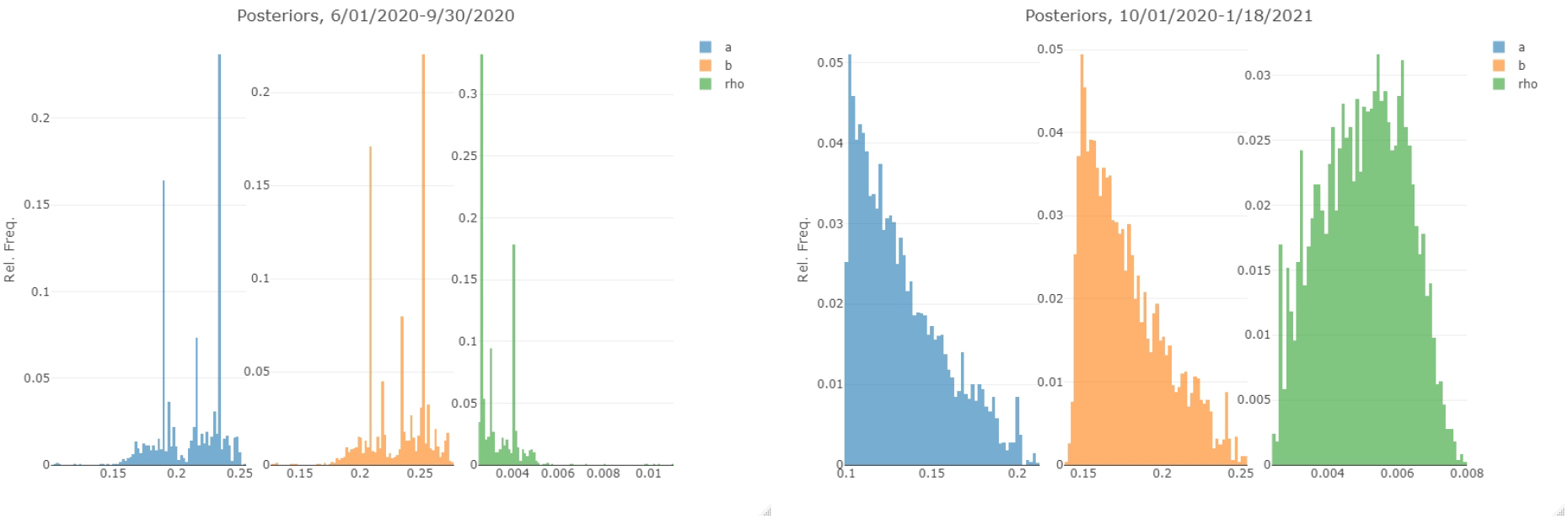
Posterior densities of the fitted parameters in the last two segments of the pandemic.

**Figure 7:**
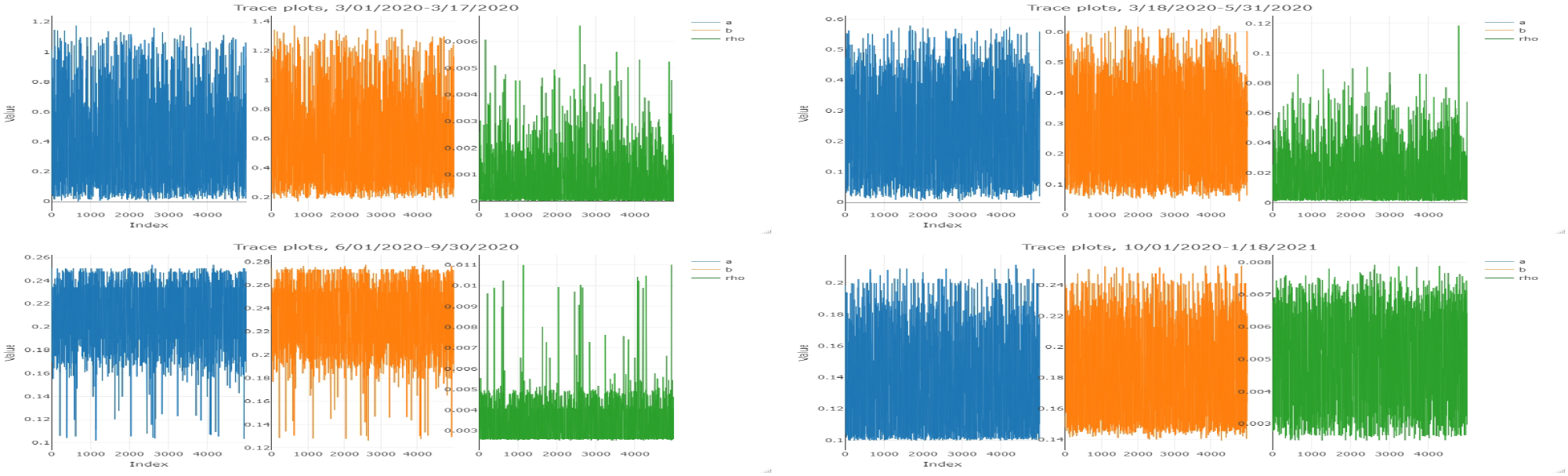
Trace plots of the fitted parameters in the four segments.

### B. Estimating Hospitalizations from Statewide Predictions of Case Numbers

Let *C*(*t*) be the trajectory of case onset times as generated by the statewide model. We wish to translate this to hospital census *h*_*i*_(*t*) for a geographic region *i*.

Consider an age group *a* ∈ 𝒜. Let *n*_*i,a*_ be the number of individuals in age group *a* in geographic region *i*, and let *n*_*i*_ = Σ_*a*∈𝒜_ *n*_*i,a*_ denote the total population size of *i*. Let *N* = Σ_*i*_ *n*_*i*_ denote the entire population size of Ohio. Converting *C*(*t*) to *h*_*i*_(*t*) involves the following steps:

i. Estimating the case onsets *c*_*i,a*_(*t*) for age group *a* in geographic region *i*.
ii. Deriving from this the onsets of severe cases *s*_*i,a*_(*t*) that will eventually require hospitalization by age group and geographic region.
iii. Using probability distributions for time from case onset to hospitalization and length of stay to estimate the hospital census *h*_*i,a*_(*t*).

#### Estimating case onsets by age group and geographic region

We consider the following factors in estimating *c*_*i,a*_(*t*) from total case onsets *C*(*t*): the population size of *i*, age structure of *i*, and the relative likelihood by age of being identified as a COVID-19 case.

We first compute the total case onsets *c*_*i*_(*t*) := Σ_*a*∈𝒜_ *c*_*i,a*_(*t*) for *i* in proportion to the population size of *i*:

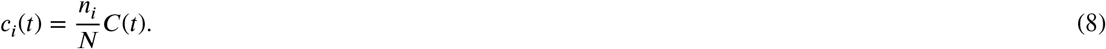

To distribute these case onsets by age, define the *relative susceptibility s*_*a*_ of age group *a* as the ratio of the proportion of observed cases in *a* relative to the proportion of the state’s population that is in *a*.

A relative susceptibility of one corresponds to the proportion of observed cases across the state matching what would be expected if cases were distributed uniformly at random across individuals. Relative susceptibilities larger than one correspond to more cases identified in *a* than would be expected at random, while relative susceptibilities less than one correspond to fewer identified cases in *a* than would be expected at random. For the Ohio data we observe far fewer cases in youth (ages less than 21) than would be expected at random (*s*_*a<*21_ ≈ 0.12), but more cases in the elderly (*s*_*a*≥85_ ≈ 1.7).

We then compute *c*_*i,a*_(*t*) by multiplying *c*_*i*_(*t*) by a probability incorporating the relative susceptibility of *a* and the age structure of *i*:

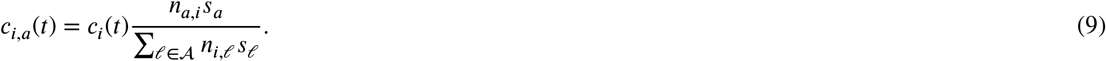

#### Onset of cases that will eventually require hospitalization

It has been documented that case outcome varies strongly with age [32]. We use the mean of the probabilities reported in [31] of severe case outcome by age group. Let *p*_*a*_ be the probability of severe case outcome for age group *a*. Then

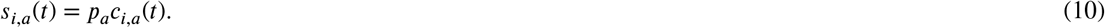

For simplicity we do not include other demographic covariates such as gender or comorbidities in the *p*_*a*_.

#### Hospital census over time

The trajectories *s*_*i,a*_(*t*) correspond to onset times of cases that will eventually require hospitalization. To translate these into hospital census of COVID-19 patients, let *w*_*a*_(*t*) be the probability distribution for time between case onset and hospitalization, and *q*_*a*_(*t*) the probability distribution for length of stay in hospital for age group *a*. Let *A*_*i,a*_(*t*) and *D*_*i,a*_(*t*) denote hospital admissions and discharges, respectively, for location *i* and age group *a* at time *t*. Then *A*_*i,a*_(*t*) corresponds to the convolution of *s*_*i,a*_ with *w*_*a*_, and *D*_*i,a*_(*t*) corresponds to the convolution of *A*_*i,a*_ with *q*_*a*_(*t*). Let *C*_*i,a*_(*t*) denote the hospital census for location *i* and age group *a* at time *t*. The difference between admissions and discharges corresponds to the change in hospital census, so integrating gives *C*_*i,a*_(*t*) :

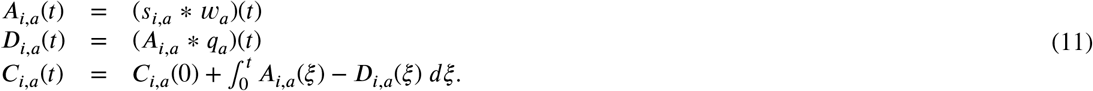

For simplicity in the above exposition we have not distinguished between hospital bed types (e.g. ICU vs. non-ICU).

## CRediT authorship contribution statement

**Wasiur R. KhudaBukhsh:** Methodology, software, writing of the manuscript. **Caleb Deen Bastian:** Methodology, software, writing of the manuscript. **Matthew Wascher:** Methodology, software, writing of the manuscript. **Colin Klaus:** Methodology, software, writing of the manuscript. **Saumya Yashmohini Sahai:** Software, writing of the manuscript. **Mark Weir:** Conceptualization of this study, methodology, writing of the manuscript. **Eben Kenah:** Conceptualization of this study, methodology, analysis, and writing of the manuscript. **Elisabeth Root:** Conceptualization of this study, methodology, writing of the manuscript. **Joseph H. Tien:** Conceptualization of this study, methodology, writing of the manuscript. **Grzegorz Rempala:** Conceptualization of this study, methodology, writing of the manuscript.

## Footnotes

1 The network parameter *κ* > 0 is defined as the ratio of network mean excess degree and mean degree, see [17] for details. It is known that *κ*= 1 corresponds to the Poisson degree network whereas *κ* > 1 corresponds to the negative binomial one.

## Notes

### Competing Interest Statement

The authors have declared no competing interest.

### Funding Statement

The study was partially supported by the National Science Foundation and National Institutes of Health under grants DMS-2027001 to EK and R01 AI116770 to GAR

### Author Declarations

The study used (or will use) ONLY openly available human data that were originally located at https://coronavirus.ohio.gov/dashboards/overview.

